# Improving Diagnostic Sensitivity for Imbalanced Musculoskeletal Disorder Data: A Sensitivity-Based Multi-Sampling Technique for Osteoarthritis Prediction

**DOI:** 10.1101/2023.11.19.23298738

**Authors:** Jun-hee Kim

**Affiliations:** Yeonsedae-gil, Maeji-ri, Heungeop-myeon, Wonju-si, Gangwon-do, 26493, Department of Physical Therapy, College of Software and Digital Healthcare Convergence, Yonsei University, Wonju, South Korea

**Keywords:** Machine learning, sample size, algorithm, sampling, musculoskeletal disorder, osteoarthritis

## Abstract

**Background:** Medical datasets containing musculoskeletal disorders may have data imbalances due to the incidence of the disease, which may limit the predictive ability, such as the sensitivity, of musculoskeletal diagnostic prediction models built from these data. This study aimed to increase the sensitivity performance of osteoarthritis (OA) prediction when building a model by adjusting an OA imbalanced dataset using a sensitivity-based multi-sampling (SMS) technique.

**Methods:** OA Data were obtained from the Korea National Health and Nutrition Examination Survey (KNHANES). SMS technique combining oversampling and undersampling was applied to the imbalanced OA data, and the RandomForest algorithm was used for machine learning modeling. Model performance was evaluated based on accuracy, sensitivity, and specificity and compared with other hybrid sampling techniques.

**Result:** In the SMS technique, ADASYN, Borderline-SMOTE, SMOTE oversampling and ENN undersampling techniques were combined and applied. The OA prediction model using the SMS technique showed the highest sensitivity (82.20) but the lowest specificity (82.26) and accuracy (82.26) compared to other hybrid models.

**Conclusion:** SMS technique offers a potential solution for improving sensitivity performance for prediction models built on medical data imbalances due to low-incidence diseases. Nonetheless, caution is warranted due to the concern that while improving sensitivity, it may decrease specificity with a trade-off.

## INTRODUCTION

Musculoskeletal diagnosis refers to the process of determining and diagnosing diseases, damage and abnormalities related to the human musculoskeletal system, which consists of structures such as muscles, bones, joints, and ligaments [1,2]. The musculoskeletal diagnosis provides medical professionals with essential information to understand the patient’s condition and to develop an appropriate treatment and care plan [1,3]. In addition, accurate musculoskeletal diagnosis allows medical personnel and related professionals to identify the extent of a patient’s illness or damage [1–3]. This allows experts to take proper care and take precautions before the disease becomes serious [3]. This accurate diagnosis can help improve the quality of life of patients who can experience pain, discomfort, and functional limitations, and speed up the recovery of daily activity [1]. Therefore, accurate diagnosis can help improve the quality of life of patients, who can experience pain, discomfort, and functional limitations, and speed up the recovery of daily activity [1,3].

The onset of musculoskeletal disorder is affected by several factors. These conditions have been reported to be primarily related to age, gender, genetic factors, lifestyle, and environmental factors [4]. Typically, lack of exercise, excessive weight, smoking, alcohol consumption, insufficient nutrient intake, excessive stress, and poor working environment can act as factors that can affect the onset of musculoskeletal disorder. Depending on these factors, the incidence of musculoskeletal disorders varies [4,5]. In 2017, the highest incidence of musculoskeletal disorders worldwide was 36.8% due to back pain, followed by osteoarthritis (OA) with 19.3%, neck pain with 18.4%, gout with 2.6%, and rheumatoid arthritis with 1.3% [5].

However, medical data, including musculoskeletal disorder data, have varying incidence rates, resulting in a data imbalance problem [6]. This data imbalance occurs when the occurrence of a particular disease is rare or uncommon [7]. Since the incidence of musculoskeletal disorders is generally lower than in healthy people, there may be fewer data points for disease classes, which may cause machine learning models to distort to primarily train normal classes, limiting their predictive ability for minority disease classes [7,8]. For this reason, data imbalances primarily seen in medical data negatively affect the performance of machine learning models [7,8]. It has been a particularly important issue in the healthcare and medical fields, and when predicting whether to diagnose a disorder with a small incidence, the sensitivity of the model may be limited due to data, which may result in missing actual patients [7,8].

Various sampling techniques are used to address data imbalance. One sampling technique is oversampling, which creates a more balanced set of data by replicating or synthesizing instances of a minority class [9,10]. Typically, oversampling technique such as SMOTE (synthetic prime oversampling technique) and ADASYN (Adaptive Synthetic Sampling) exist [9,10]. Undersampling, on the other hand, involves reducing the number of instances in multiple classes, such as Tomek link or Neighbourhood-based undersampling [11,12]. Also, Combinations of oversampling and undersampling techniques can also be used to maximize the benefits of both approaches. SMOTE-Tomek, which combines SMOTE and Tomek, or SMOTE-ENN, which combines SMOTE and Edited Nearest Neighbors, can be seen as examples of hybrid sampling [13,14]. These techniques are effective at mitigating data imbalances but can lead to overfitting or potentially losing valuable information when training machine learning models [9–11,13,15].

The purpose of this study was to develop and apply sampling techniques to improve the low sensitivity performance of models built with medical datasets such as musculoskeletal disorder that indicate data imbalances according to low incidence rates. Therefore, in this study, a sensitivity-based multiple sampling (SMS) technique was developed, and the SMS technique was applied to construct a dataset with an emphasis on improving sensitivity to predict OA, a musculoskeletal disorder with a low incidence. And this technique was applied to machine learning model training and compared to the performance of machine learning models of other sampling techniques.

## MATERIALS AND METHODS

### Data Sources

The flowchart of this study is shown in Figure 1. The study samples were obtained from Korea National Health and Nutrition Examination Survey (KNHANES), a national periodic survey conducted by the Korea Disease Control and Prevention Agency. This study used 8110 datasets, the first-year data of the 8th KNHANES (2019–2020). Among the datasets, data that meet the following criteria were selected. 1) Participants over the age of 19 2) Data with diagnosis as OA 3) Data with no missing values in variables to be used as predictors. Of the 8110 samples without data missing values, 621 samples with only osteoarthritis without rheumatoid arthritis and 4988 data samples without both OA and rheumatoid arthritis were finally selected for this study. Variables representing individual characteristics, physical activity, lifestyle, and descriptions used in this study are shown in Table 1.

**Table 1.** List of variables and variable descriptions (mean + SD)

|  | **OA (621)** | **Normal (4988)** | **Description** |
| --- | --- | --- | --- |
| Sex | M:116 / F:505 | M:2403 / F:2585 | Gender (M: male / F: female) |
| Age | 67.84 ± 9.95 | 49.04 ± 16.21 | Age (year) |
| Height | 156.72 ± 8.27 | 164.74 ± 9.06 | Height (cm) |
| Weight | 61.35 ± 10.70 | 64.95 ± 12.97 | Weight (kg) |
| BMI | 24.92 ± 3.46 | 23.82 ± 3.61 | Weight/Height<sup>2</sup> |
| BO1_1 | 1:426 / 2:90 / 3:105 | 1:3122 / 2:671 / 3:1195 | Weight change (1: none / 2: increase / 3: decrease) |
| HE_wc | 87.69 ± 9.46 | 83.51 ± 10.48 | Waist circumference (cm) |
| GS | 21.97 ± 7.68 | 29.53 ± 9.91 | Grip strength (kg) |
| LQ4_00 | 1:131 / 2:490 | 1:278 / 2:4710 | Activity limitation (1: yes / 2: no) |
| EQ5D | 0.86 ± 0.15 | 0.96 ± 0.09 | Quality of life (EQ5D index) |
| EC1_1 | 1:245 / 2:376 | 1:3199 / 2:1789 | Work (1: yes / 2: no) |
| EC_wht_23 | 14.91 ± 21.17 | 27.91 ± 21.92 | Work time |
| BE3_71 | 1:2 / 2:619 | 1:82 / 2:4906 | Work activity (1: yes / 2: no) |
| BE3_75 | 1:11 / 2:610 | 1:558 / 2:4430 | Leisure activity (1: yes / 2: no) |
| BD3_2 | 1.97 ± 1.78 | 3.00 ± 1.74 | Smoking (per day) |
| BD1_11 | 0.72 ± 3.44 | 2.42 ± 5.89 | Drinking alcohol (per week) |
| BP16_1,<br>BP16_2 | 6.39 ± 1.56 | 6.88 ± 1.28 | Sleep time (hour) |

**Figure 1.**
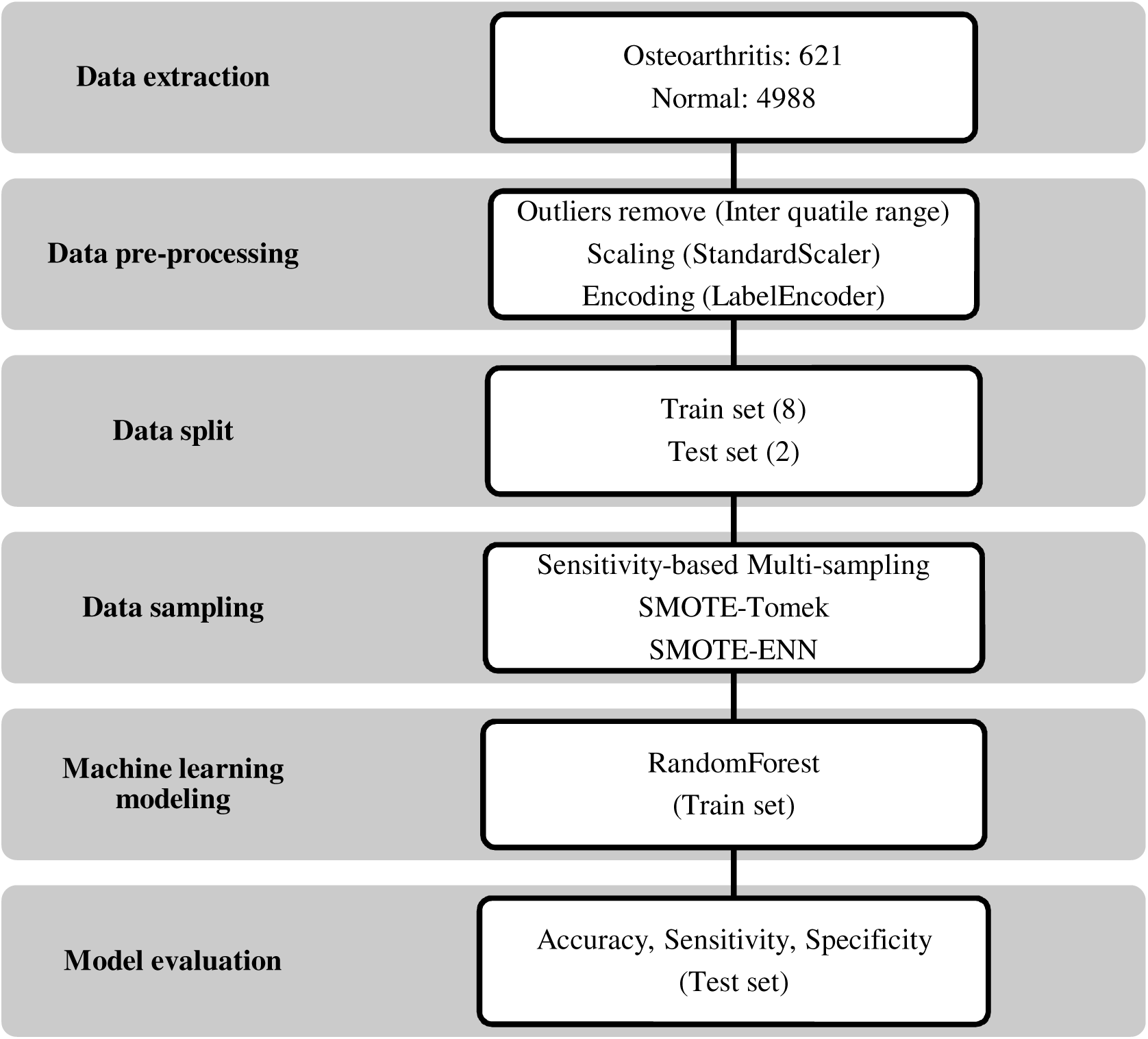
Flowchart of this study

### Data Pre-processing

Inter Quarter Range (IQR) and Boxplot were utilized to detect and eliminate outliers. Outliers in numerical data were identified and processed based on IQR, and scaling was performed using the StandardScaler function for numerical variables. Categorical variables were converted into dummy data using the LabelEncoder function. This pre-processed data was divided into training and test data at an 8:2 ratio and utilized for model learning and performance evaluation.

### Sensitivity-based Multi-sampling and Hybrid Sampling

SMS, which was designed in this study, is a technique that is a combination of oversampling and undersampling techniques (Figure 2). According to the results of the test performance evaluation of each model learned with data sampled by five oversampling techniques, the top three oversampling techniques were selected based on sensitivity score. In addition, the undersampling technique with the highest sensitivity score was selected according to the test performance evaluation results of each model learned with data sampled using the four undersampling techniques. In the original data, the data was augmented by dividing half the sample size difference between classes into three selected oversampling techniques. And finally, a selected undersampling technique reduced the data by half of the sample size difference between classes. Hybrid sampling techniques, SMOTE-Tomek and SMOTE-ENN, were employed for comparison with the SMS technique in this study. Hybrid sampling involves a combination of oversampling and undersampling techniques to harness the benefits of both approaches.

**Figure 2.**
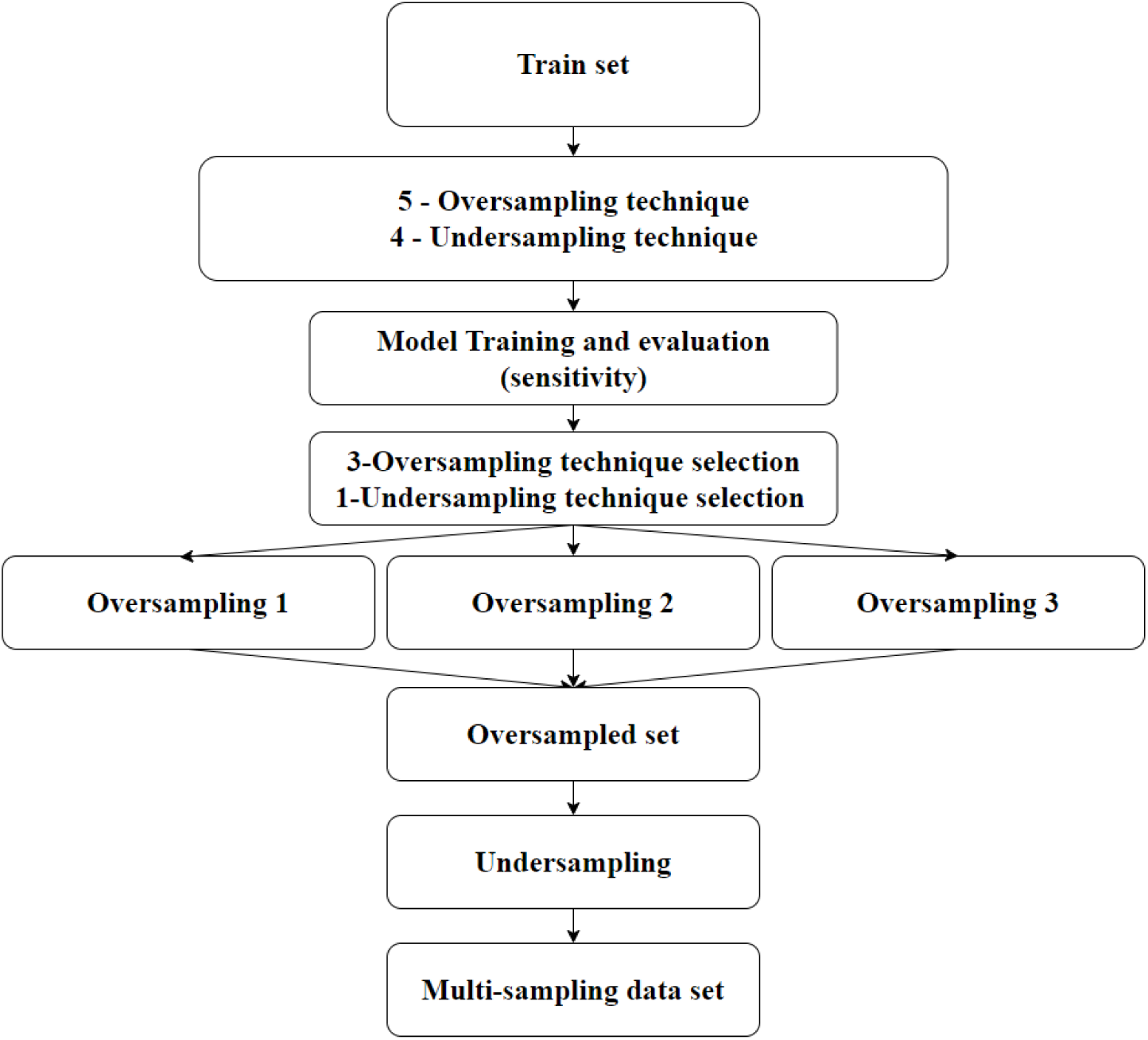
Sensitivity-based Multi-sampling algorithm

### Machine-learning modeling

In this study, the RandomForest machine learning algorithm was used to compare the differences in OA prediction performance between sampling techniques. Hyperparameters were not fine-tuned and default values were used, and the default values were as follows: n_estimators (decision-making number of trees): 100, criterion (splitting criterion): “gini”, max_depth (maximum tree depth): no limit, min_samples_split (minimum number of samples for node splitting): 2, min_samples_leaf (minimum number of samples required from a terminal node) : 1, max_features (maximum number of features to use for splitting at each node): auto. The default values of hyperparameters were used to create a RandomForest machine-learning model for OA diagnosis prediction.

### Model evaluation

In applying the SMS technique, the performance of machine learning models built by each oversampling and undersampling technique evaluated accuracy, sensitivity, and specificity. In addition, the performance of machine learning models built with respective training datasets sampled by SMS, SMOTE-Tomek, and SMOTE-ENN was also evaluated and compared for accuracy, sensitivity, and specificity.

## RESULTS

### Oversampling and Undersampling model performance

The accuracy of RandomForrest models built with training data applied with oversampling techniques was found to be 89.61 high from 87.90 (Table 2). In terms of sensitivity, ADASYN, Borderline-SMOTE, and SMOTE were found to have high performance in order.

**Table 2.** Oversampling techniques application and model evaluation scores.

|  | Accuracy | Sensitivity | Specificity |
| --- | --- | --- | --- |
| **SMOTE** | 88.71 | 59.32 | 92.18 |
| **ADASYN** | 87.90 | 60.17 | 91.18 |
| **Borderline-SMOTE** | 88.62 | 60.17 | 91.98 |
| **SVMSMOTE** | 89.61 | 57.63 | 93.39 |
| **Synthetic Over-sampling** | 88.08 | 57.63 | 91.68 |

The accuracy of RandomForrest models built with training data applied with this downsampling technique was found to be 89.52 high at 86.74 (Table 3). In terms of sensitivity, the model built by the sampling technique of Repeated Edited Nearest Neighbours showed the highest performance at 69.49.

**Table 3.**
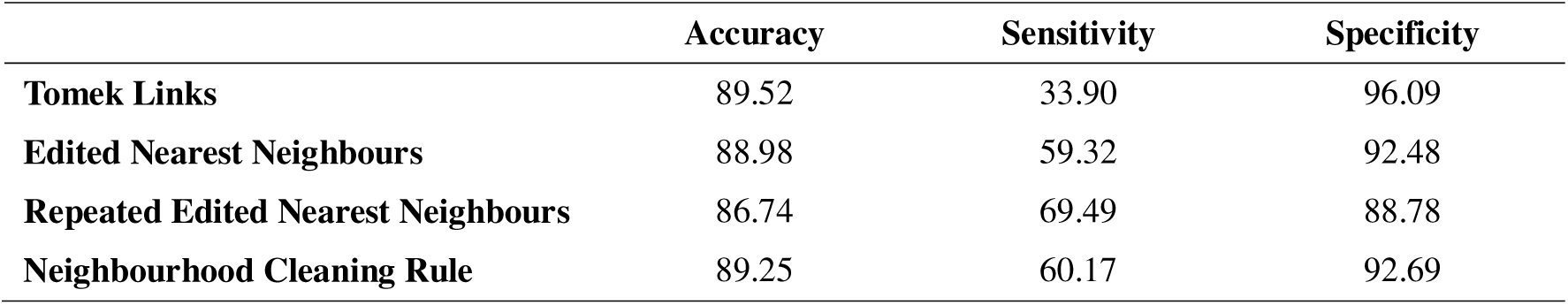
Undersampling techniques application and model evaluation scores.

### Sensitivity-based Multi-sampling

The three oversampling techniques chosen by SMS were ADASYN, Borderline-SMOTE, and SMOTE, and the undersampling technique was Repeated Edited Nearest Neighbours (Figure 3). A total of 1758 OA data were added to the original train data, 586 each by three oversampling techniques according to the SMS algorithm. Then, 1247 normal data were removed by Repeated Edited Nearest Neighbours, selected by the under-sampling technique, leaving only 2743 normal data.

**Figure 3.**
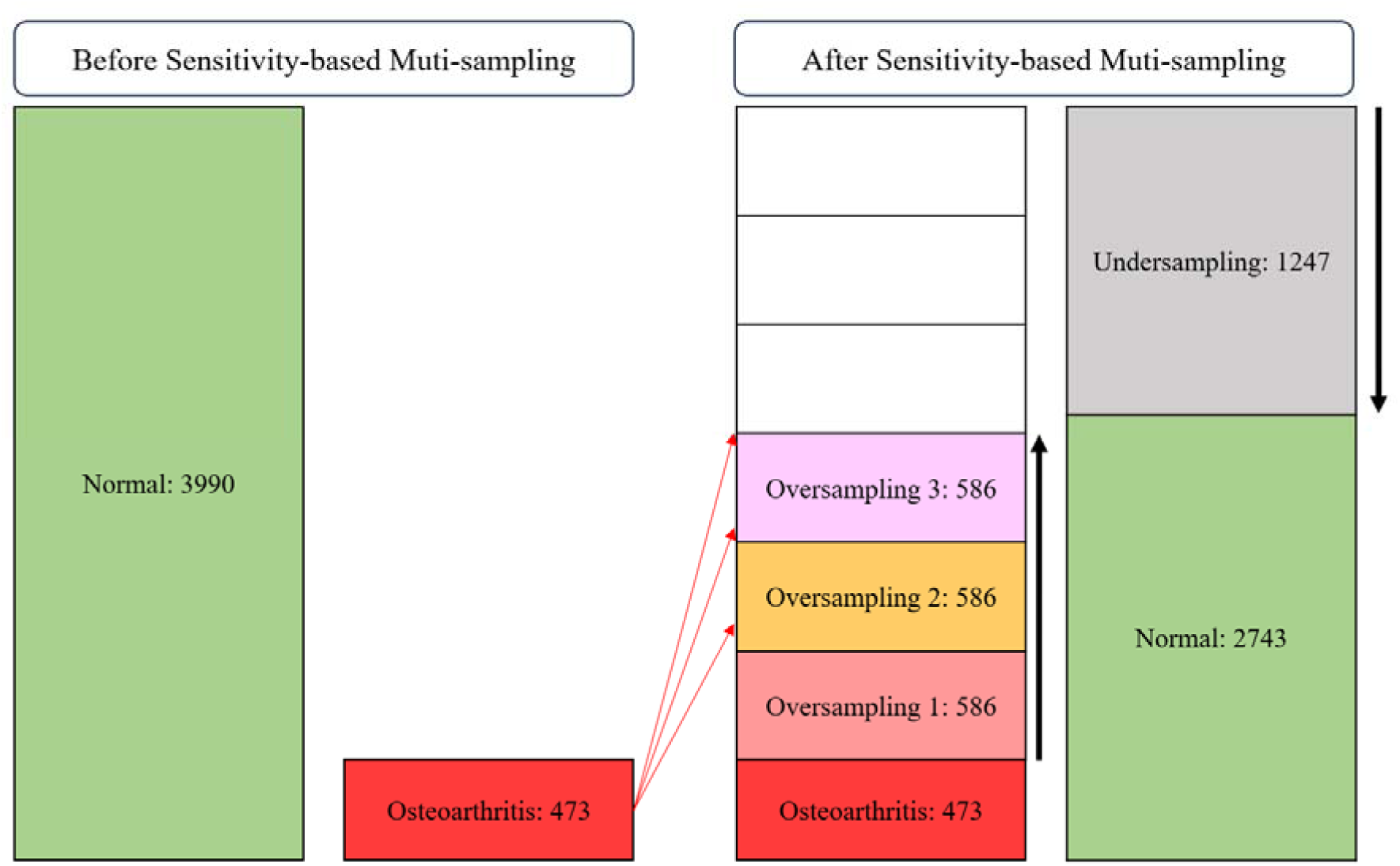
Train set structure after Sensitivity-based Multi-sampling technique

### Sensitivity-based Multi-sampling and Hybrid Sampling Model Performance

Table 4 shows the performance of the RandomForrest model for OA prediction built with train datasets sampled by the Hybrid sampling techniques. The accuracy performance of the model built using the SMS technique was 82.26, the lowest among the three models, but the sensitivity performance was the highest at 82.20. On the other hand, the accuracy performance of the model built using the SMOTE-Tomek technique was found to be the highest at 88.35, but the lowest sensitivity performance of 56.78. SMOTE-ENN showed the second highest performance with accuracy performance of 84.05 and sensitivity of 77.12, respectively. In terms of specificity, SMOTE-TOMEK was the highest at 92.08, SMOTE-ENN was second at 84.87, and SMS was the lowest at 82.26.

**Table 4.** Hybrid sampling techniques application and model evaluation score.

|  | **Accuracy** | **Sensitivity** | **Specificity** |
| --- | --- | --- | --- |
| **Sensitivity-based Multi-sampling** | 82.26 | 82.20 | 82.26 |
| **SMOTE-Tomek** | 88.35 | 56.78 | 92.08 |
| **SMOTE-ENN** | 84.05 | 77.12 | 84.87 |

## DISCUSSION

In this study, a SMS technique was introduced and applied to address the data imbalance problem in musculoskeletal disorder prediction, especially focusing on OA. An OA prediction model using the SMS technique was constructed, and sensitivity performance was compared with other prediction models. The model was constructed by applying a combination of oversampling (ADASYN, Borderline-SMOTE, SMOTE) and undersampling (iteratively edited nearest neighbors) using a SMS technique. The accuracy and specificity of the built model decreased compared to other models, but in terms of sensitivity, it showed higher performance than other models.

In medical data, where data imbalance inevitably occurs due to the rate of disease occurrence, model performance often deteriorates due to data imbalance [16,17]. If a prediction model is created with imbalanced medical data, sensitivity performance may decrease [16,17]. Shim et al. (2020) built an osteoporosis risk prediction model using osteoporosis patients and normal subjects with a sample size that was approximately three times that of the patients [17]. The sample ratio differed by only about 3 times, but when the model was built using the RandomForest algorithm, the sensitivity was only 59 [17]. In the case of medical data, various sampling techniques have been proposed to improve the poor performance caused by data imbalance [7,18]. Sampling techniques to resolve data imbalance are largely divided into undersampling and oversampling [15,19]. Oversampling involves creating additional instances of minority classes to balance the class distribution [15,19]. This helps the model learn minority classes better, but generating too many synthetic examples can lead to overfitting. This means that the model may become too tailored to the training data and perform poorly on new data [15,19]. Undersampling involves reducing the number of instances of a majority class, which can lead to information loss issues [15,19]. This results in the loss of valuable information present in the majority class, potentially resulting in a less representative model [15,19]. If many classes contain important patterns or variations, removing instances may reduce the model’s ability to generalize to unseen data [15,19].

To address the problems associated with oversampling and undersampling applied to building models containing such imbalanced medical data, various hybrid sampling techniques have been proposed and used. Zeng et al. (2016) proposed a preprocessing technique combining SMOTE with Tomek link and applied it to imbalanced medical data sets of three diseases [20]. The combined technique of SMOTE and Tomek Link showed marked improvement in 31, 27, and 30 out of 32 evaluation indicators for diabetes, Parkinson’s disease, and spinal disease compared to the oversampling using SMOTE alone [20]. Lamari et al. (2021) used an ensemble technique that combines SMOTE, which synthesizes samples belonging to minority classes, and Edited Nearest Neighbors (ENN), which examines the neighbors of a given sample and removes unnecessary samples [14]. Using ensemble techniques, they improved the sensitivity performance of appendicitis, diabetes, and Parkinson’s disease classification models from an average of 78 to 82 [14]. Khushi et al. (2021) built a model by applying both hybrid sampling techniques (SMOTE-ENN, SMOTE-Tomek) to the lung cancer dataset, and the AUC performance of this model increased from 50 to 65.50 [16]. Our study also showed performance improvement in terms of sensitivity. With only oversampling or undersampling, sensitivity performance ranged from 33.90 to 69.49. In the case of hybrid techniques combining oversampling and undersampling, sensitivity performance ranged from 56.78 to 77.12, except for SMS. In comparison, SMS showed a high sensitivity performance of 82.20.

Sensitivity is a measure of how accurately true positives can be detected among positive cases and is very important in diagnosing a disease [21–23]. In other words, the higher the sensitivity, the fewer false negatives, allowing more effective identification of actual positive cases [21]. Especially in disease diagnosis, missing a positive test can be a big problem, and if a positive test is not accurately detected, the patient may not receive appropriate treatment or measures, which can lead to serious problems due to delayed diagnosis or inaccurate results [22]. Considering the aspect of sensitivity in medical diagnosis, this study designed a sampling technique to improve low sensitivity performance due to data imbalance. The reason why SMS had the highest sensitivity in this study may be because oversampling and undersampling techniques were selected in order of high sensitivity. However, there is a trade-off between sensitivity and specificity [24,25]. When sensitivity is high, the model is good at finding positive samples without missing them, but this makes it easier for the model to predict positive ones and may mispredict negative ones as positive [24]. This may result in an increase in the false positive rate and a decrease in specificity [24]. In other words, sensitivity and specificity are complementary indicators, and improving one may decrease the other, and in medical diagnosis, sensitivity may be more important than specificity [26–28]. For this reason, sensitivity-based multi-sampling technique to improve sensitivity would have reduced specificity and further reduced accuracy.

This study has some limitations. First, the effectiveness of the SMS technique may vary depending on the characteristics of the disease dataset used. Different disease datasets may have different patterns of imbalance depending on the incidence of the disease, so sampling rates may need to be adjusted. Second, the hyperparameters for the RandomForest machine learning algorithm were not tuned and default values were used.

Since hyperparameter tuning can improve model performance, it will be necessary to investigate the impact of hyperparameter tuning on sensitivity-based multi-sampling technique. Third, since the variables used to construct this OA prediction model were not medical measurement variables used to diagnose OA, it is unclear how reliably they can be applied in actual medical settings. Therefore, although this model can assist in diagnosing OA, it cannot replace medical diagnostic criteria.

## CONCLUSION

This study attempted to solve the decrease in sensitivity performance caused by data imbalance in predicting musculoskeletal disorder, focusing on OA, by introducing and applying the SMS technique. In this study, the RandomForest machine learning algorithm was used, and the performance of the SMS technique was compared with other techniques combining oversampling and undersampling. According to the study results, the SMS technique, which was designed to improve the sensitivity of predicting OA, had the highest sensitivity, but showed lower specificity, which is a trade-off, compared to other models. In conclusion, SMS technique provides valuable insight into the challenge of addressing data imbalances in musculoskeletal disorder prediction. SMS technique has shown potential to improve sensitivity, an important indicator of disease diagnosis, but their use should be approached cautiously, considering the trade-offs and limitations observed in specificity and overall model performance.

## Data Availability

This study used data samples obtained from the 8th Korean National Health and Nutrition Survey (KNHANES) of the Korea Centers for Disease Control and Prevention from 2019 to 2020.

